# A school-based cluster-randomized pragmatic trial to control dental caries using minimally invasive interventions

**DOI:** 10.1101/2024.10.22.24315936

**Authors:** Ryan Richard Ruff, Aditi Ashish Gawande, Qianhui Xu, Tamarinda J. Barry Godín

**Author notes:** Corresponding author: Ryan R. Ruff.

## Abstract

**Background:** Evidence-based non-surgical interventions to halt the progression of dental caries, the most prevalent noncommunicable disease in the world, include atraumatic restorative treatment (ART) and silver diamine fluoride (SDF). Data are needed on their effectiveness when used in school caries prevention programs.

**Methods:** In this school-based, cluster-randomized pragmatic trial conducted from February 1, 2018 to June 1, 2023, 48 primary schools in New York City were randomly assigned to receive either silver diamine fluoride or atraumatic restorations for untreated caries on any mesial, occlusal, distal, buccal, and lingual surface of permanent molars, premolars, and primary molars. All children then received fluoride varnish. Children were treated by either dental hygienists, pediatric dentists, or medical nurses (SDF group only). Dental caries was diagnosed as any lesion scoring either 5 or 6 on the ICDAS scale. The primary outcome was the caries control rate.

**Results:** A total of 7418 children were enrolled in the trial, of which 1668 (861 in the SDF group, 807 in the ART group) presented with treatable dental caries and completed at least one follow-up observation. The total surface-level failure in the SDF group was 38.3%, compared to 45.5% in the ART group. There were 2167 failures observed in SDF participants over 1372 person-years, compared to 2116 ART failures over 1291 person-years, yielding an incidence rate ratio of 0.96 (95% CI = 0.86, 1.03). At the subject level, 45.5% of SDF recipients experienced at least one surface failure, compared to 53.3% of ART recipients. There were no significant differences in the risk of recurrent surface failure between treatments (HR = 0.92, 95% CI = 0.82, 1.04).

**Conclusions:** In this four-year pragmatic trial of school-based utilization of minimally invasive interventions for dental caries, similar control rates were observed in children receiving either SDF or ART. These results support the use of secondary preventive therapies for school dental programs.

**Author roles:** RRR conceptualization, formal analysis, methodology, funding acquisition, supervision, writing-original draft; AG: formal analysis; QX: formal analysis; TJBG: data curation, investigation.

## 1 Introduction

Dental caries (tooth decay) is a worldwide public health crisis, afflicting billions of children and adults [1] who often lack access to effective preventive or therapeutic care [2]. It is also highly inequitable, as those from low-income or minority families shoulder a disproportionate burden of disease [3]. Untreated caries may increase the risk of systemic noncommunicable diseases including cancer, diabetes mellitus, cardiovascular diseases, and neurodegenerative conditions [4]. Caries also affects child development, reducing educational performance [5] and oral health-related quality of life [6], and is attributed to over thirty million hours of lost seat time in schools per year [7].

Integrating preventive and therapeutic dental services into schools can increase access to care, reduce the risk of caries, and may improve educational performance [8]. The U.S. Department of Health and Human Services’ Community Preventive Services Task Force recommends school sealant programs to prevent dental caries [9], and the Centers for Disease Control and Prevention funds school-based sealant programs (SBSPs) in multiple states. These programs are both clinically and cost-effective [10; 11]. However, managing existing caries in children who are unlikely to seek out traditional, office-based care remains a critical issue in dental public health.

Silver diamine fluoride (SDF) and glass ionomer atraumatic restorative treatment (atraumatic restorations, or ART) are minimally invasive interventions that can effectively arrest or control caries, and are included on the World Health Organizations’ Model List of Essential Medicines. SDF is a topical solution that inhibits the growth of cariogenic bacteria and contributes to the remineralization of enamel and dentine caries [12], whereas atraumatic restorative treatment removes decalcified tissue with hand instruments before applying adhesive fillings to restore the cavity [13]. Silver diamine fluoride is estimated to arrest anywhere from 47 to 90 percent of caries lesions after one application [14] and is considered to be a practical, affordable approach to community caries prevention, particularly in low-socioeconomic areas [15]. In contrast, the average annual failure rate for single- and multiple-surface ART restorations in primary molars is estimated to be 5 and 17%, respectively [16], though this may be outperformed by conventional restorations [17]. Like SDF, prior studies of ART in community settings conclude that it is acceptable and effective in socioeconomically deprived groups [18].

The CariedAway study was a pragmatic randomized trial that assessed SDF and ART for the control of dental caries when used in a school-based oral health program [19]. In this paper, we estimate the overall and comparative effects of treatment on caries control over four years. A secondary objective was to determine whether posterior application of silver diamine fluoride resulted in subsequent anterior caries control.

## 2 Methods

CariedAway was a longitudinal, cluster-randomized pragmatic trial conducted from February 1, 2018 to June 1, 2023 in eligible elementary schools in New York, NY, USA [20]. The study received IRB approval from the New York University School of Medicine and is registered at www.clinicaltrials.gov (#NCT03442309). CariedAway results are reported following the Enhancing the Quality and Transparency of Health Research (EQUATOR) guidelines.

### Participants

Any school in the New York City geographic area with a student population consisting of at least 50% black and/or Hispanic/Latino and at least 80% receiving free and reduced lunch was eligible to participate. These inclusion criteria were used as they typically have the highest burden of disease in the New York metropolitan area. Amongst participating in schools, any child with parental informed consent and child assent was enrolled in the study. While care was provided to any child meeting these criteria, analysis was restricted to those aged 5-13 years.

### Randomization

Schools were block randomized to either the experimental or active control condition using a random number generator performed by RRR and verified by TBG. All children in a school received the same treatment.

### Interventions and Procedures

For untreated caries, study participants received either silver diamine fluoride followed by fluoride varnish (to mask the bitter aftertaste of SDF) or glass ionomer cement (GIC) atraumatic restorative treatment followed by fluoride varnish. In the experimental group, a 38% concentration SDF solution (2.24 F-ion mg/dose) was applied to any posterior, asymptomatic, cavitated lesions using a microbrush. In the active control group, placement of GIC atraumatic restorations were applied to all frank, asymptomatic, cavitated lesions. Additionally, dental sealants were applied to sound dentition in the active control, but were not included in analysis. Treatments were provided in a private, dedicated room in a school using portable dental mirrors, explorers, and dental chairs with a frontal light source. All treatments were administered by either dental hygienists (SDF and ART), a supervising pediatric dentist (SDF or ART), or registered nurses (SDF) under patient-specific standing orders signed by a supervising dentist. Examination and treatment were provided biannually, except for disruptions in care due to COVID-19.

### Data Collection and Diagnosis Protocol

Demographic data including age, sex, and race/ethnicity were obtained prior to examination from informed consent documents. Caries diagnosis was performed following a visual-tactile dental screening according to the International Caries Detection and Assessment System (ICDAS) adapted criteria in epidemiology and clinical research settings [21]. Any lesion with an ICDAS score of 5 (distinct cavity with visible dentine) or 6 (extensive/more than half the surface distinct cavity with visible dentine) was recorded as caries. At each observational visit, the first instance of dental caries was recorded and treated following the described clinical protocols. SDF was applied at each observational visit regardless of surface caries status, while ART was only reapplied if failed.

### Outcome Definition

Caries control was determined if a carious surface (mesial, occlusal, distal, buccal, and lingual) on treatable teeth (permanent molars, premolars, and primary molars) received the assigned treatment and did not present with recurrent caries at any subsequent observation over the course of the study. Any caries recurrence was classified as control failure, and any surfaces on which the first observation of caries occurred at the subjects’ last study visit were removed from analysis. The per-person control rate

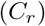

was then computed as the number of controlled surfaces divided by the total number of treated surfaces, and the per-person failure rate was computed as

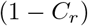

The time elapsed for each controlled or failed surface from initial treatment was then recorded.

### Power

As caries control over four years was not a primary endpoint of the CariedAway trial, an a priori power analysis was not performed.

### Statistical Analysis

Descriptive statistics for the full study enrollment and for those participants with at least one follow-up observation and at least one carious surface were computed overall and by treatment group (means, standard deviations, and frequencies). We calculated the total number of controlled and failed surfaces observed for each study participant over a maximum of four years of follow-up, estimating the incidence rate ratio of failure. Controlled and failed surfaces were also determined by sex and race/ethnicity. To account for potential within-school correlation, we subsequently analyzed subject-level failure rates using binomial regression with cluster adjustment. To determine the effect of disease severity on failure rates we included a predictor for the per-person number of carious surfaces and an interaction effect between disease severity and treatment. We then considered the possibility of recurrent surface failure using frailty models with predictors for treatment type and selected participant demographic variables. A random effect was included for dependence among recurrent event times. For simplicity we first analyzed data ignoring higher-level school clustering, but then used nested frailty models to include random effects at individual and school levels. Finally to determine the association between posterior SDF application and anterior surface caries control, we restricted the sample to those participants assigned to the silver diamine fluoride group that presented with both posterior and anterior decay on any surface at baseline. We then calculated the number of decayed anterior surfaces at baseline and number of controlled surfaces at first follow-up and measured the strength of the association using Pearson’s correlation. Statistical analysis was conducted in R v.4.3.3. Statistical significance was determined at p*<*0.05.

## 3 Results

A total of 7418 participants were enrolled in the CariedAway trial, consisting of 3739 (50.4%) randomized to the silver diamine fluoride group and 3679 (49.6%) randomized to the atraumatic restoration group (Table 1, Figure 1). The average baseline decay prevalence was 26.7%, and the average age at baseline was 7.6 years (SD=1.9). Over 65% of enrolled participants reported being from either Hispanic/Latino or black race/ethnicity. For the analytic sample (participants with at least one follow-up observation and presented with at least one surface-level carious lesion prior to their last treatment visit), 1668 met inclusion criteria consisting of 861 (51.6%) randomized to the SDF group and 807 (48.4%) randomized to the ART group. Approximately 68% of the analytic sample had initial caries at baseline, while the remaining 32% developed caries over the course of the study. The analytic sample was equally distributed with respect to baseline decay, sex, age, and race/ethnicity.

**Table 1:**
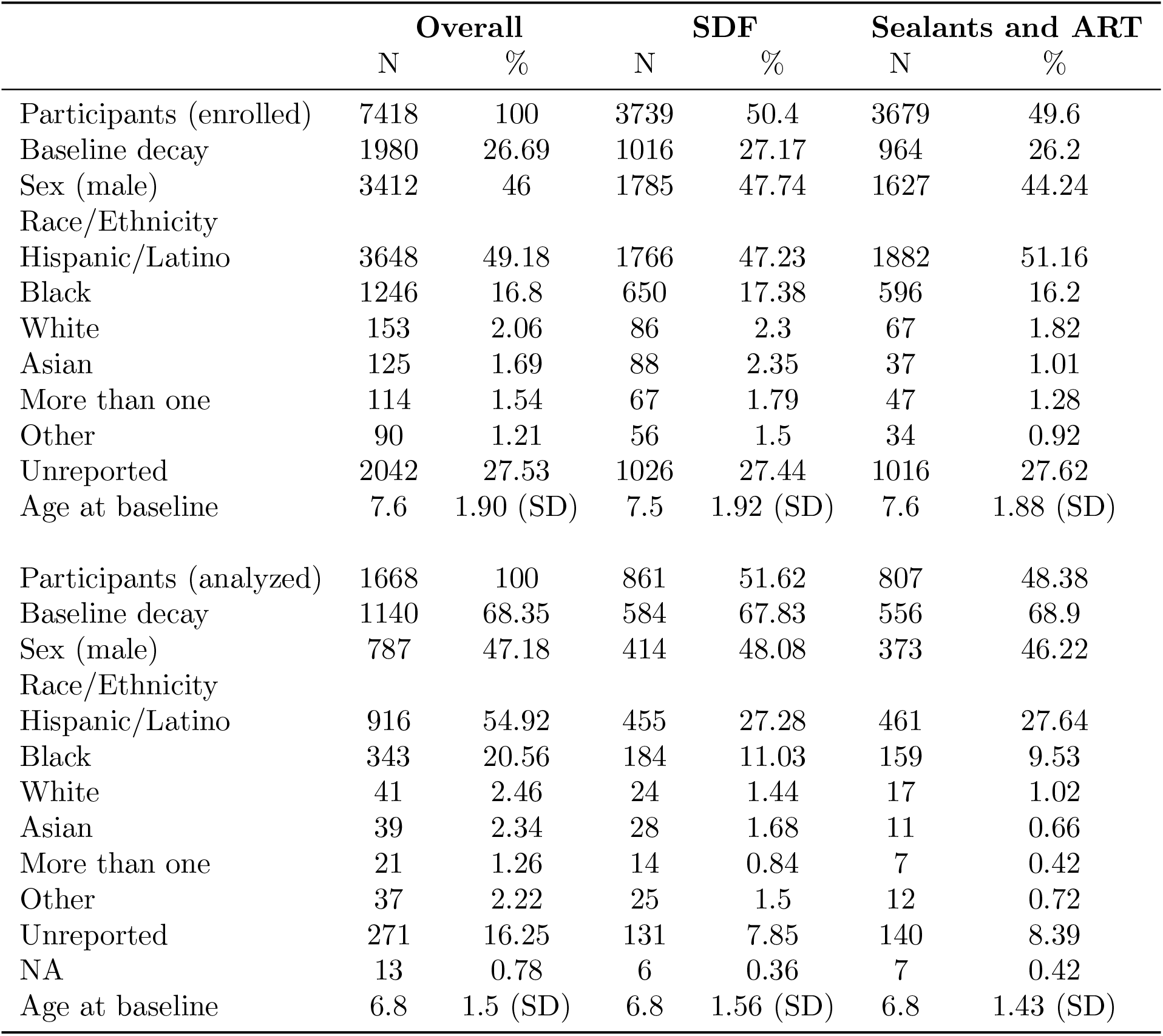
Patient characteristics for total enrollment and analytic sample.

|  | **Overall** |  | **SDF** |  | **Sealants and ART** |  |
| --- | --- | --- | --- | --- | --- | --- |
|  | N | % | N | % | N | % |
| Participants (enrolled) | 7418 | 100 | 3739 | 50.4 | 3679 | 49.6 |
| Baseline decay | 1980 | 26.69 | 1016 | 27.17 | 964 | 26.2 |
| Sex (male) | 3412 | 46 | 1785 | 47.74 | 1627 | 44.24 |
| Race/Ethnicity |  |  |  |  |  |  |
| Hispanic/Latino | 3648 | 49.18 | 1766 | 47.23 | 1882 | 51.16 |
| Black | 1246 | 16.8 | 650 | 17.38 | 596 | 16.2 |
| White | 153 | 2.06 | 86 | 2.3 | 67 | 1.82 |
| Asian | 125 | 1.69 | 88 | 2.35 | 37 | 1.01 |
| More than one | 114 | 1.54 | 67 | 1.79 | 47 | 1.28 |
| Other | 90 | 1.21 | 56 | 1.5 | 34 | 0.92 |
| Unreported | 2042 | 27.53 | 1026 | 27.44 | 1016 | 27.62 |
| Age at baseline | 7.6 | 1.90 (SD) | 7.5 | 1.92 (SD) | 7.6 | 1.88 (SD) |
| Participants (analyzed) | 1668 | 100 | 861 | 51.62 | 807 | 48.38 |
| Baseline decay | 1140 | 68.35 | 584 | 67.83 | 556 | 68.9 |
| Sex (male) | 787 | 47.18 | 414 | 48.08 | 373 | 46.22 |
| Race/Ethnicity |  |  |  |  |  |  |
| Hispanic/Latino | 916 | 54.92 | 455 | 27.28 | 461 | 27.64 |
| Black | 343 | 20.56 | 184 | 11.03 | 159 | 9.53 |
| White | 41 | 2.46 | 24 | 1.44 | 17 | 1.02 |
| Asian | 39 | 2.34 | 28 | 1.68 | 11 | 0.66 |
| More than one | 21 | 1.26 | 14 | 0.84 | 7 | 0.42 |
| Other | 37 | 2.22 | 25 | 1.5 | 12 | 0.72 |
| Unreported | 271 | 16.25 | 131 | 7.85 | 140 | 8.39 |
| NA | 13 | 0.78 | 6 | 0.36 | 7 | 0.42 |
| Age at baseline | 6.8 | 1.5 (SD) | 6.8 | 1.56 (SD) | 6.8 | 1.43 (SD) |

**Figure 1:**
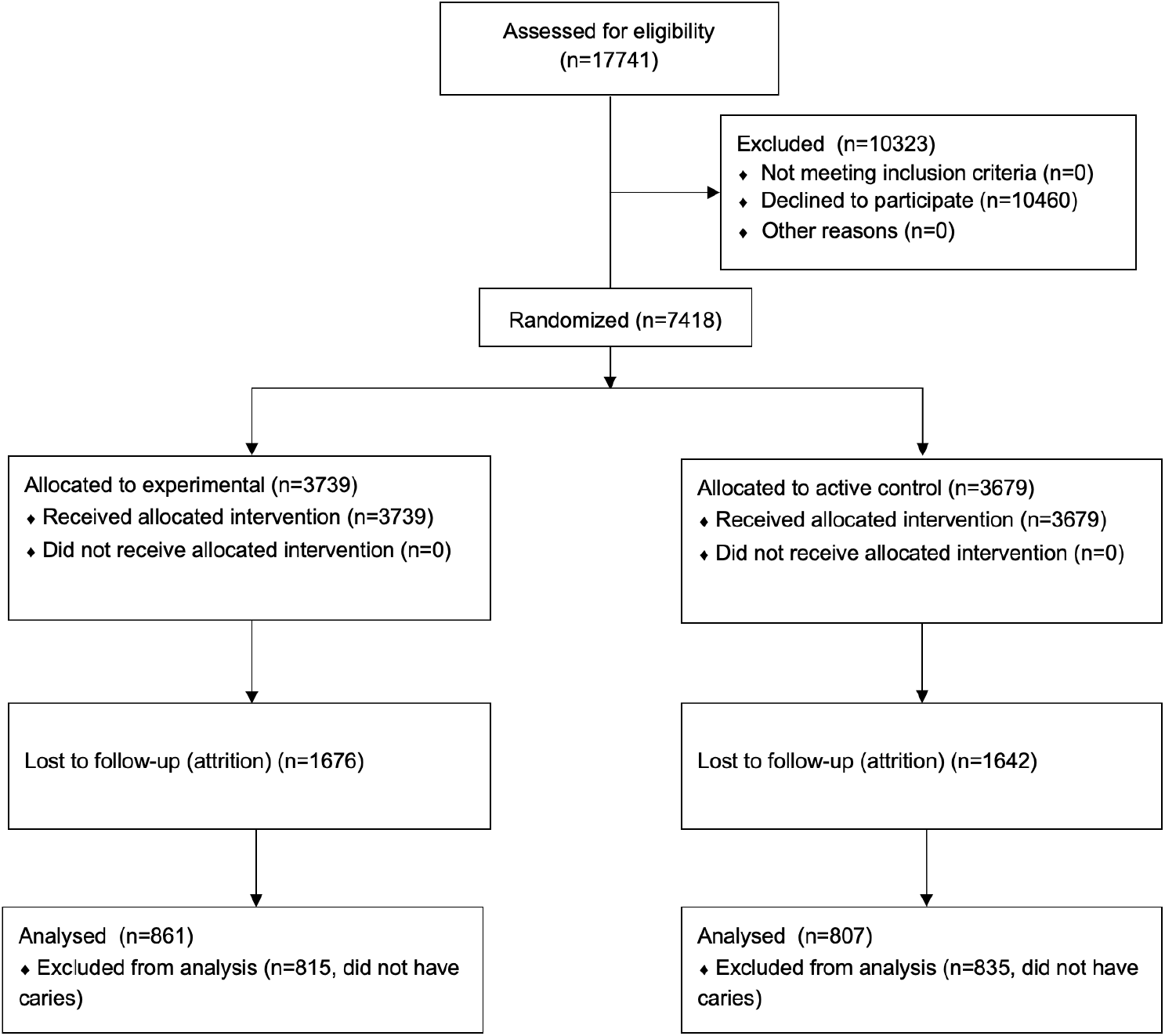
Study enrollment flowchart.

The distribution of surface-level caries in the analytic sample (Figure 2) indicate that the majority of participants had between one and six surfaces with decay (e.g., 314 subjects presented with one carious surface, 494 subjects with two carious surfaces, and 258 subjects with three carious surfaces). Within the SDF group, there were 5651 total carious surfaces that received treatment (Table 2). Of these surfaces, 2167 presented with subsequent decay, for a surface-level failure rate of of 38%. In the ART group, 4647 total carious surfaces were treated, of which 2116 presented with later decay, for a failure rate of 45.5%. The person-level failure rates in both groups were slightly higher, estimated as 46% and 53% for SDF and ART groups, respectively. When considering the total time of observation (1371 person-years in the SDF group, 1291 person-years in the ART group), the incidence rate ratio was 0.96 (95% CI = 0.91, 1.02, p = 0.226). Similar rates were found across treatments by sex, but there were significant differences between treatments in select racial/ethnic groups.

**Table 2:**
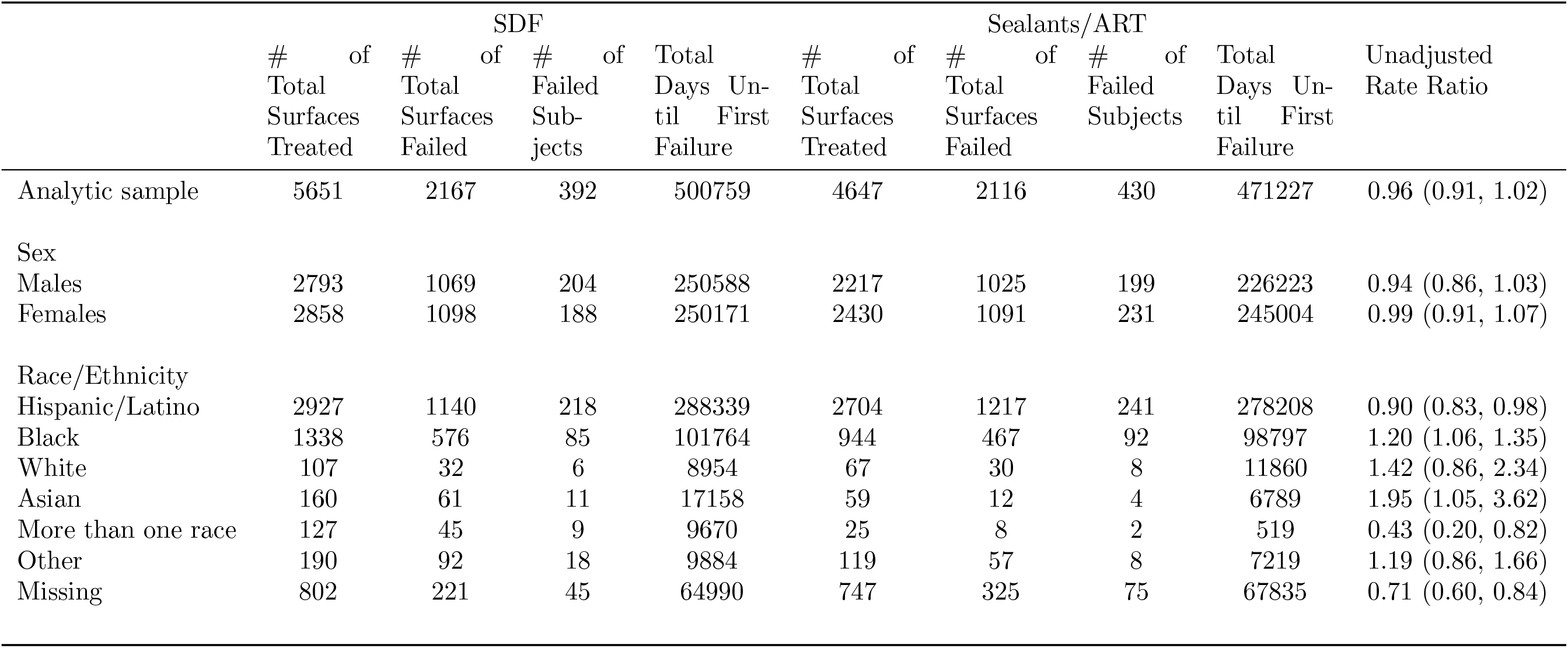
Unadjusted rate ratio of any surface-level control failure.

**Figure 2:**
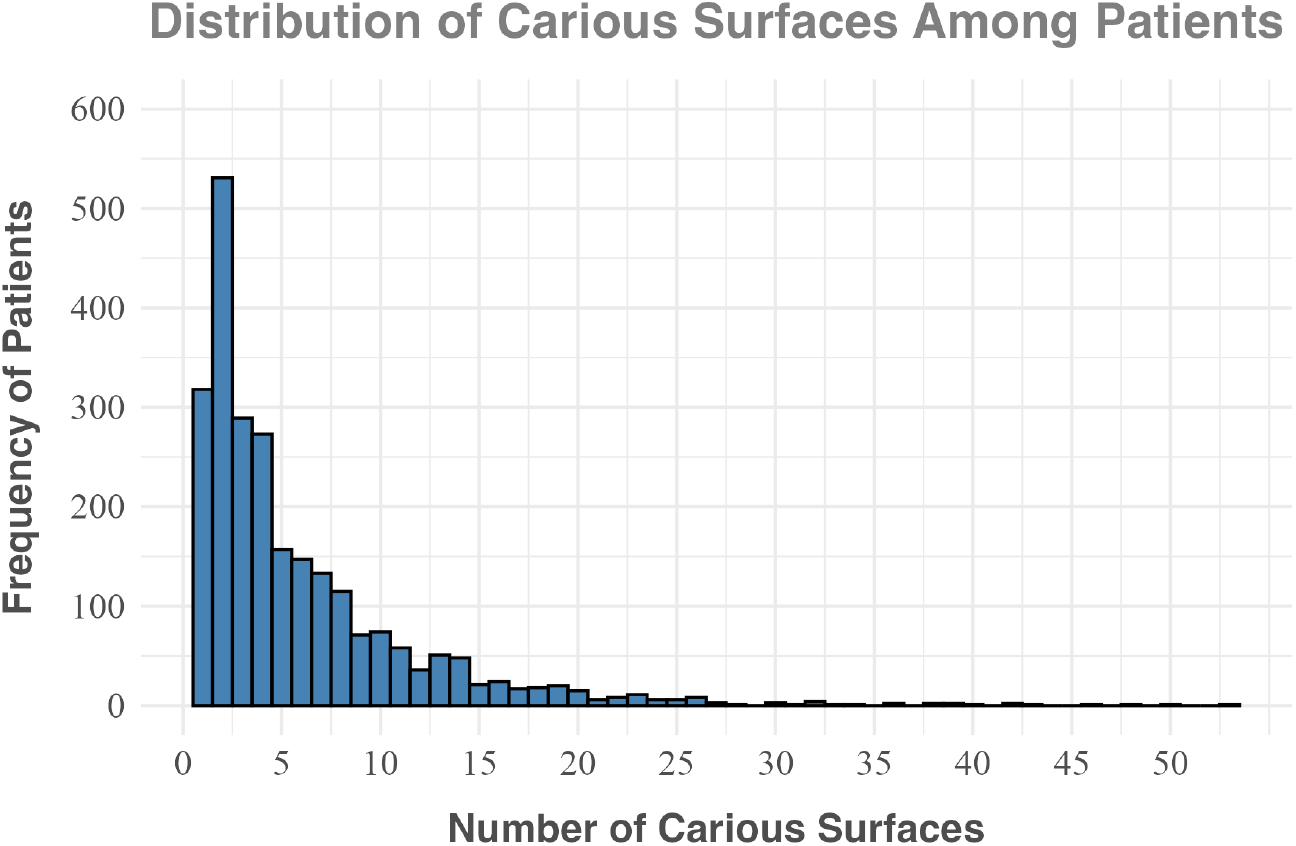
Histogram of disease severity in the analytic sample.

After adjusting for the potential clustering within school (Table 3), the odds of person-level failure was significantly reduced in children receiving silver diamine fluoride (OR = 0.64, 95% CI = 0.42, 0.97). Disease severity was significantly related to control failure, with each additional decayed surface increasing the odds of failure by approximately 20% (OR = 1.20, 95% CI = 1.13, 1.27). However, there was no significant interaction between treatment and disease severity (OR = 0.96, 95% CI = 0.90, 1.02).

**Table 3:** Individual level failure by treatment and disease severity.

|  | **lnOdds** | **SE** | **OR** | **95% CI L** | **95% CI U** | **p-value** |
| --- | --- | --- | --- | --- | --- | --- |
| SDF (vs ART) | -0.449 | 0.211 | 0.63827 | 0.4218945 | 0.9656054 | 0.034 |
| Disease Severity | 0.18 | 0.031 | 1.19722 | 1.1263699 | 1.2737942 | 0.0001 |
| Interaction | -0.044 | 0.033 | 0.95695 | 0.8967304 | 1.0222438 | 0.19 |

Similar to unadjusted rate ratios, children receiving SDF had a slight reduction (8%) in the recurrent risk of surface failure (Table 4) conditional on unmeasured heterogeneity. The reduction was not statistically significant (95% CI = 0.82, 1.04). In nested models, the effects were not appreciably different (HR = 0.82, p = 0.232) and there were no significant differences between sex and race/ethnicity.

**Table 4:** Hazard ratios of surface caries control failure in CariedAway trial participants (analytic sample)

|  | **HR** | **95% CI L** | **95% CI U** |
| --- | --- | --- | --- |
| SDF (vs ART) | 0.92 | 0.82 | 1.04 |
| Males | 1.05 | 0.93 | 1.19 |
| Race/Ethnicity |  |  |  |
| Black | 1.11 | 0.95 | 1.3 |
| White | 0.69 | 0.46 | 1.03 |
| Asian | 0.96 | 0.6 | 1.55 |
| Multiple | 1.54 | 0.9 | 2.65 |
| Other | 1.11 | 0.75 | 1.66 |
| Missing | 1.09 | 0.9 | 1.31 |

Finally, there were 462 anterior surfaces with decay at baseline across 108 participants that had both posterior and anterior decay and received silver diamine fluoride on posterior carious lesions. In these participants, 920 posterior teeth received SDF. At the first follow-up observation, six anterior surfaces were arrested (0.013%) across two subjects. There was no correlation between anterior surface arrest and the number of posterior teeth treated with SDF (r = -0.001, p = 0.991).

## 4 Discussion

The CariedAway study was a school-based clinical trial of minimally invasive treatments for caries management. Over 1600 children received either silver diamine fluoride or atraumatic restorations on more than 10,000 carious tooth surfaces. The surface- and participant-level failures rates were 38.3 and 45.5% in the SDF group and 45.5 and 53.3% in the ART group, respectively. SDF recipients outperformed ART ones at the individual level, but there were no significant differences in the risk of recurrent failure in adjusted analyses.

While the use of SDF and ART in United States schools is limited, prior school-based application of atraumatic restorations reported reductions in the per-visit risk of caries by approximately 20% in children with untreated decay [22]. We previously reported that a single application of silver diamine fluoride or ART halted caries progression in 56% and 46% of high-risk schoolchildren two years after initial treatment, respectively, [23]. Our results support similar conclusions for repeat application after four years. Additionally, previous estimates were computed at the individual-level, while our data are based on more robust surface-level failure rates. Results also corroborate international studies, where SDF has been provided as part of a preschool oral health program in Hong Kong, demonstrating both clinical effectiveness and a high rate of acceptance among parents [15]. Similarly, an ART program in Hong Kong kindergartens reported 30-month restoration survival rates ranging from 79% to 33% [24], depending on restoration classification, and one in Brazilian public schools estimated a six-month success rate of approximately 82% [25]. Outside of school settings, both silver fluoride and ART were found to be effective in a community trial in Australian Aboriginal children [26], and a mobile dental program operating in underserved Mexican communities reported a 66% retention rate for atraumatic restorations after two years [27].

The effectiveness of using SDF and ART for caries management in schools may be modified by the severity of disease, such as size and depth of the carious lesion. Differences in arrest attributed to treatment may also be affected by disease severity [28]. The American Academy of Pediatric Dentistry (AAPD) reports a wide range of caries arrest rates for SDF, with effectiveness varying by cavity size and tooth location [14], and notes the arresting capacity of SDF may be greater in less severe lesions. For ART, a systematic review of atraumatic restorative treatment in children estimates a 71% and 67% success rate after one and two years, respectively, but the success of multiple surface restorations was significantly lower than single-surface ones [27]. Accordingly, the AAPD policy reports that single surface applications have higher survival than multi-surface restorations [29]. Caries diagnosis in the present study was based on an ICDAS score of 5 or 6, but we were unable to stratify by severity in analysis. While we do present evidence that the risk of failure increases with each additional diseased surface, this may be a result of having more surfaces at risk rather than a proxy for disease severity, and more research is needed.

Frequency of application is also likely to impact caries control. A review of SDF studies suggests semiannual application [30] to mitigate the reduction in effectiveness for caries arrest that may occur over time, while the AAPD Clinical Practice Guidelines recommends a single application with follow-up after two to four weeks, reapplying where indicated [14]. Other approaches use more intensive early application, such as weekly application over a three week period, in an attempt to enhance arresting effects [31]. Similar to other school-based dental programs [32], the clinical protocols for CariedAway stipulated semiannual application of both SDF and ART (if not retained, in the case of atraumatic restorations). However, this initial schedule was disrupted due to school closings related to the COVID-19 pandemic. As a result, the elapsed time between treatments had considerable variability for enrolled participants. Our results suggest that observed failure rates with repeat application are similar to those after a single application [23]. It may be the case that the effectiveness of SDF and ART is reduced somewhat in pragmatic settings, such as school-based caries prevention. Regardless, the overall success rate is encouraging for community caries management.

The direct effects of SDF has received considerable study [12; 33; 34]: the mechanistic action of silver diamine fluoride includes reducing the growth of cariogenic bacteria through the antibacterial effect of silver, and also by promoting the remineralization of enamel and dentine caries [12]. Silver ions in SDF are thought to prevent bacterial aggregation through reaction with bacterial cellular surface proteins [35], harden soft carious lesions through reaction with phosphate or chloride ions resulting in the formation of silver salts (e.g., silver chloride), and increase the alkalinity of the environment through the formation of ammonium compounds hypothesized to have an acid-buffering effect [36]. However, the potential indirect effects of application on adjacent caries activity has not been meaningfully examined. SDF application in CariedAway was restricted to posterior dentition to mitigate any negative impact that SDF staining might have on facial aesthetics and oral health-related quality of life. We subsequently explored whether posterior application controlled anterior lesions and if there was any dose-response relationship. Our data do not indicate any such association and we observed no correlation between the number of posterior surfaces treated and any subsequent anterior arrest.

There are a number of study limitations in addition to those already discussed. Due to the gaps in care due to COVID-19, exfoliation of primary dentition in the intervening period may have occurred. As a result, follow-up for affected tooth surfaces would stop if recorded as missing, and then resume for newly erupted permanent dentition if later diagnosed to be carious. Additionally, it is possible that if a study participant were to visit an outside dental provider, the examining dentist may put a permanent restoration in place of any dentition previously treated with SDF or ART. While we consider this to be unlikely given the target population enrolled, any observation of a filling on a surface previously treated with either SDF or ART would not be recorded as decayed. This would inflate the success percentage of treatments.

## 5 Conclusions

Both silver diamine fluoride and atraumatic restorative treatment are effective in the longitudinal control of dental caries when used in school-based prevention. Incorporating these and other minimally invasive techniques can help improve access to critical preventive oral medicine and decrease health inequities.

## Data Availability

Data will be available upon request and completion of a data use agreement by 12/1/2025

